# Identification and validation of tolerogenic dendritic cells-related biomarkers in diabetic retinopathy

**DOI:** 10.1101/2025.10.01.25337114

**Authors:** Wenwang Liang, Songman Li, Lingjuan Liu, Junpeng Huang, Xiaohui Lai, Lijuan Yang, Chunyi Wei

**Author notes:** These authors contributed equally to this work (Wenwang Liang, Songman Li).

## Abstract

**Objective:** Diabetic retinopathy (DR) is a primary microvascular complication of diabetes. Its pathogenesis is associated with chronic inflammation and immune responses. While tolerogenic dendritic cells (tolDCs) are critical for suppressing excessive inflammation and maintaining immune homeostasis, their function in DR is not well characterized. This study aims to identify tolDCs-related biomarkers and elucidate their underlying mechanisms in DR through integrated bioinformatics and clinical validation.

**Methods:** All data used in this study were obtained from public databases. Biomarkers were identified through differential analysis, machine learning, and expression validation. Subsequently, enrichment analysis and immune infiltration were used to investigate the functional mechanisms of the biomarkers. Finally, clinical peripheral blood specimens were obtained for validation using reverse transcription-quantitative PCR (RT-qPCR).

**Results:** A total of 2,096 differentially expressed genes (DEGs)1 were identified between DR and control groups. A total of 6,267 tolDCs-related genes were identified. After that, 51 key genes were selected. A total of 2 biomarkers, TFEC and CHMP2A, were identified through machine learning and expression validation selecting. GSEA results demonstrated TFEC enrichment in 994 pathways, among which was “Cytoplasmic Ribosomal Proteins.” CHMP2A was enriched in 748 pathways, including “tRNA Processing.” The pathways enriched by both TFEC and CHMP2A included “apoptosis,” “VEGFA-VEGFR2 signaling,” and “signaling by VEGF,” among others. A total of 8 differential immune cell types were identified, including resting mast cells. TFEC showed the strongest correlation with activated NK cells (correlation coefficient (cor) = -0.49, *p*< 0.001), CHMP2A with resting mast cells (cor = 0.49, *p*< 0.001). RT-qPCR analysis revealed a significant upregulation of TFEC and downregulation of CHMP2A in the DR group, aligning with the bioinformatics findings.

**Conclusion:** TFEC and CHMP2A were identified as biomarkers and were involved in the development of DR through tolDCs.

## Introduction

Diabetic retinopathy (DR) is a leading blinding complication of diabetes, affecting approximately 30–40% of diabetic patients and contributing to visual impairment or blindness in over 100 million people worldwide [1]. Diabetic macular edema is a leading cause of central vision loss in these patients. As an immune-privileged tissue, the retina is protected by a highly sophisticated system that prevents systemic immune attack. Hyperglycemia-induced vascular changes disrupt the blood-retinal barrier, triggering a cascade of macular edema. Low-grade, chronic inflammatory activation further promotes severe retinal damage and chronic maculopathy [2]. Substantial evidence indicates that chronic inflammation and immune dysregulation are closely associated with the pathogenesis of DR. Current understanding of DR pathology includes microangiopathy, neurodegeneration, and mild to moderate inflammatory responses—a state also referred to as microinflammation [3]. This microinflammation represents a low-grade, persistent systemic inflammatory state, and thus DR is considered a chronic low-grade inflammatory disease. Inflammation plays a central role throughout DR progression, manifested by increased expression of systemic and ocular inflammatory molecules such as elevated C-reactive protein, increased neutrophil count, and upregulation of intraocular inflammatory biomarkers [4, 5]. In both DR animal models and clinical patients, vascular endothelial cells upregulate molecules that mediate leukocyte adhesion, leading to leukostasis in the retinal and choroidal microcirculation [6]. Currently, intravitreal anti-vascular endothelial growth factor (anti-VEGF) agents and long-acting hormones are primary treatments to inhibit or delay DR progression. However, a significant proportion of patients exhibit poor or no response to these intravitreal therapies, ultimately progressing to blindness and imposing substantial psychological and economic burdens on patients, their families and society [7]. In recent years, numerous studies have explored inflammatory biomarkers as objective indicators to evaluate the relationship between systemic inflammation and diabetic complications [8]. The systemic immune-inflammatory index is a systemic inflammatory marker that accurately assesses inflammatory and immune response status [9]. It provides a comprehensive measure of a patient’s immune and inflammatory status, demonstrating potential for early prediction and guidance in the treatment of diabetic complications.

Tolerogenic Dendritic Cells (tolDCs) are a key type of cell in the field of immune regulation and possess the capacity to produce diverse immunomodulatory cytokines and metabolites, subsequently altering the local microenvironment to promote the differentiation of leukocytes (including T and B lymphocytes) into immunosuppressive phenotypes or maintain immune homeostasis. Many experiments have confirmed that tolDCs can produce Interleukin-10 (IL-10), which can induce non-responsiveness in effector and memory CD4+ T cells [10]. TolDCs inhibit the development of autoimmune-mediated hyperglycemia into insulin-dependent type 1 diabetes mellitus, while preserving a critical mass of β cells capable of restoring normal blood glucose levels to some extent in newly diagnosed clinical disease [10]. Clinical studies have shown that subcutaneous injection of tolDCs can effectively suppress antigen-specific pro-inflammatory T cells in patients with type 1 diabetes, leading to the generation of regulatory T cells and IL-10, thereby intervening in the progression of the disease [11]. However, the molecular mechanisms between tolDCs and DR remain to be further explored.

This study uses data from public databases on DR and tolDCs, using integrated approaches combining differential expression profiling, computational modeling, and molecular validation, we sought to discover tolDCs-related biomarkers in DR pathogenesis. It aims to understand the predictive value of these biomarkers for DR, their biological functions, and their associations with the immune microenvironment, potentially offering novel therapeutic avenues for DR intervention.

## 2 Materials and methods

### 2.1 Data acquisition

All gene expression data analyzed in this DR investigation were obtained from the GEO database (Accessed on February 28th, 2025). The GSE221521 dataset (platform GPL24676) was designated as the training cohort, comprising 69 DR patient peripheral blood samples and 50 control samples, with other available samples excluded from analysis. The GSE185011 (GPL24676) dataset was used as the validation set, the dataset comprised matched cohorts of 5 DR patients and 5 healthy controls, all analyzed using peripheral blood mononuclear cells (PBMCs), with the other samples excluded. The GSE23371 (GPL570), GSE52894 (GPL10558), GSE182528 (GPL570), and GSE56017 datasets were used as datasets related to tolDCs. In the GSE23371 dataset, 3 activated tolerogenic dendritic cell samples (tolDCs group) and 3 tolerogenic precursor dendritic cell samples (control group) were selected, with the remaining samples excluded. In the GSE52894 dataset, 5 lipopolysaccharide-treated tolDCs samples (tolDCs group) and 4 tolerogenic precursor dendritic cell samples (control group) were selected, with the other samples excluded. In the GSE182528 dataset, 5 tolDCs samples derived from healthy donor in vitro (tolDCs group) and 5 monocyte-derived dendritic cell samples were selected (control group), with the remaining samples excluded. In the GSE56017 dataset, 8 tolerogenic monocyte-derived dendritic cell samples (tolDCs group) and 7 mature monocyte-derived dendritic cell samples (control group) were selected, with the other samples excluded.

### 2.2 Differentially expressed genes (DEGs) analysis

The DESeq2 package (v 1.38.0) [12] was utilized to identify DEGs1 between the DR and control groups in the GSE221521 dataset (adj. *p* < 0.05 and |log_2_ FC| >0.5). The ggplot2 package (v3.4.1) was used to visualize DEGs1 through a volcano plot [13], annotating it to highlight the top five most significantly upregulated and downregulated genes. The ComplexHeatmap package (v 2.15.1) [14] was employed to construct a heatmap representation of the 10 most differentially expressed genes in both directions. Differential analysis between tolDCs and control groups was conducted on the GSE23371, GSE52894, GSE182528, and GSE56017 datasets using the limma package (v3.56.2) [15] (*p* < 0.05 and |log_2_ FC| >0.5). The methods for generating the volcano plot and heatmap were the same as described above. The resulting genes were respectively named DEGs2, DEGs3, DEGs4, and DEGs5. Subsequently, DEGs2, DEGs3, DEGs4, and DEGs5 were merged, and duplicate genes were removed to obtain the tolDCs-related genes.

### 2.3 Key genes acquisition

The ggVenn package (v 0.1.10) [16] was used to integrate the DEGs1 with tolDCs-related genes. The obtained genes were designated as candidate genes. Based on candidate genes, the clusterProfiler package (v4.2.2) was employed to conduct comprehensive functional enrichment analysis of the gene sets [17], including GO (*p* < 0.05) and KEGG enrichments (*p* < 0.05). The results displayed the top 10 enriched terms and top 5 KEGG pathways, selected based on their respective gene counts. We generated a PPI network for candidate genes through STRING (Search Tool for Retrieval of Interacting Genes/Proteins), retaining only high-confidence interactions (interaction score > 0.15). The result was visualized using Cytoscape (v 3.10.2) [18]. Subsequently, the candidate genes were analyzed using five algorithms from CytoHubba: BottleNeck, Closeness, Radiality, Betweenness, and Stress. The consensus among the top 100 genes across the five algorithms was graphically represented using an upset plot visualization. The resulting genes were designated as key genes.

### 2.4 Biomarkers acquisition

The glmnet package (v4.1.8) was employed to conduct LASSO regression analysis with optimal lambda selection [19] on key genes across all samples in the GSE221521 dataset, where genes that were not penalised to 0 were considered as feature genes 1. Additionally, the randomForest package (v 4.7.1.2) [20] was used to perform the Random Forest algorithm. When the 5-fold cross-validation error rate reached its minimum, the importance of each gene was recorded. The most biologically significant genes were identified as feature genes based on their importance scores. Candidate biomarkers were identified by finding the overlapping genes between feature sets 1 and 2, with results displayed via ggVennDiagram (v0.1.10). Differential expression of candidate biomarkers was assessed using Wilcoxon rank-sum tests (*p* < 0.05) in both GSE221521 and GSE185011 datasets. Only genes showing consistent expression patterns (up/down-regulation) in both cohorts were ultimately validated as biomarkers. Additionally, a ROC curve was generated for the GSE221521 dataset using the pROC package (v1.18.5) [21], and the AUC value was calculated to evaluate the predictive performance of the biomarkers (AUC > 0.7).

### 2.5 Gene set enrichment analysis (GSEA)

The reference gene set was obtained from the “c2.cp.v2024.1.Hs.symbols.gmt” file in MSigDB. Pathway enrichment analysis was conducted using the clusterProfiler package (v4.2.2) through GSEA methodology. The psych R package (v2.2.9) was employed to calculate comprehensive correlation matrices comparing each biomarker’s expression profile against all genes in GSE221521 [22]. The resulting correlation coefficients were ranked and these ranked lists were subjected to thresholding at |NES| > 1 and *p* < 0.05 to generate biomarker-associated gene lists. The top 5 pathways were presented based on *P*-value.

### 2.6 Immune microenvironment analysis

Immune cell infiltration profiles were quantified using CIBERSORT to estimate proportions of 22 leukocyte subsets in GSE221521, excluding samples that failed to meet the significance threshold (*p* > 0.05). The Wilcoxon rank-sum test (*p* < 0.05) was employed to identify statistically significant variations in immune cell proportions between DR patients and controls. Spearman correlation analysis was performed on all GSE221521 samples to explore the relationships between biomarkers and differential immune cell types, as well as between the immune cell types themselves, using the psych package (v 2.2.9) (|cor| > 0.30, *p* < 0.05).

### 2.7 Reverse transcription-quantitative PCR (RT-qPCR)

This study enrolled a total of 10 volunteers, comprising 5 DR patients and 5 matched controls, who were recruited by the same attending physician at our hospital in January 2025. Peripheral blood samples were collected from all participants, and the expression levels of tolDCs-related biomarkers were experimentally validated using reverse transcription-quantitative polymerase chain reaction (RT-qPCR). Written informed consent was obtained from all individuals, and the study protocol was approved by the Ethics Committee of Ruikang Hospital Affiliated to Guangxi University of Chinese Medicine (Ethics Approval No. KY2024-213).

The inclusion criteria for the DR experimental group were as follows: (1) diagnosis of type 2 diabetes mellitus; (2) confirmation of DR by fundus photography, optical coherence tomography (OCT), or fluorescein angiography; (3) age between 45 and 85 years. The exclusion criteria for all participants were: (1) presence of severe systemic diseases (e.g., renal failure with an estimated glomerular filtration rate (eGFR) < 60mL/min/1.73m², malignancy, or active immune diseases); (2) congenital or hereditary ocular diseases (e.g., congenital cataract, retinitis pigmentosa, or congenital/developmental glaucoma); (3) acute or progressive infectious eye diseases (e.g., infectious endophthalmitis or keratitis); (4) history of traumatic ocular diseases (e.g., traumatic cataract or ocular rupture); (5) history of ocular surgery within the past 3 months (e.g., cataract extraction, vitrectomy, or intravitreal anti-VEGF injection); (6) significant opacities of the refractive media that could affect the quality and assessment of fundus imaging.

Total RNA of 10 samples was extracted using TRIzol (Ambion, USA). The extracted total RNA was reverse transcribed into cDNA using the SweScript First Strand cDNA Synthesis Kit (Servicebio, Wuhan, China) following manufacturer’s protocols. RT-qPCR analysis was conducted using Servicebio’s 2× Universal Blue SYBR Green qPCR Master Mix (Wuhan, China) under standard cycling conditions.

The primer sequences used for PCR amplification are provided in **Supplementary Table 1**. GAPDH served as the endogenous control gene for normalization of expression data. Quantitative PCR amplification was performed using the CFX Connect Real-Time PCR Detection System (Bio-Rad, USA) under the following cycling parameters: 1 min at 95℃ for pre-denaturation, 20 s at 95℃ for denaturation, 20 s at 55℃ for annealing, and 30 s at 72℃ for extension, for a total of 40 cycles. mRNA levels were quantified relative to the reference gene using the 2^-ΔΔCT^ method [23].

### 2.8 Data analysis

Statistical analysis was implemented in R (v 4.2.2), with between-group differences evaluated via Wilcoxon testing (significance defined as *p* < 0.05).

## 3. Results

### 3.1 DEGs acquisition

Analysis of the GSE221521 dataset revealed 2,096 differentially expressed genes (DEGs) in DR samples, comprising 1,567 upregulated and 529 downregulated transcripts (Fig. 1A). The GSE23371 cohort exhibited 4,021 significant DEGs in tolDCs compared to controls, with 1,653 genes showing increased expression and 2,368 genes demonstrating decreased expression (Fig. 1B). Analysis of the GSE52894 dataset revealed 3,285 differentially expressed genes (DEGs) in tolDCs, consisting of 1,448 upregulated and 1,837 downregulated transcripts (Fig. 1C). Analysis of the GSE182528 dataset identified 419 differentially expressed genes (DEGs) in tolDCs, including 270 upregulated and 149 downregulated transcripts (Fig. 1D). The GSE56017 cohort demonstrated 1,346 significant DEGs in tolDCs relative to controls, with 860 genes exhibiting increased expression and 486 genes showing decreased expression (Fig. 1E). Finally, 6,267 tolDCs-related genes were identified.

**FIGURE 1.**
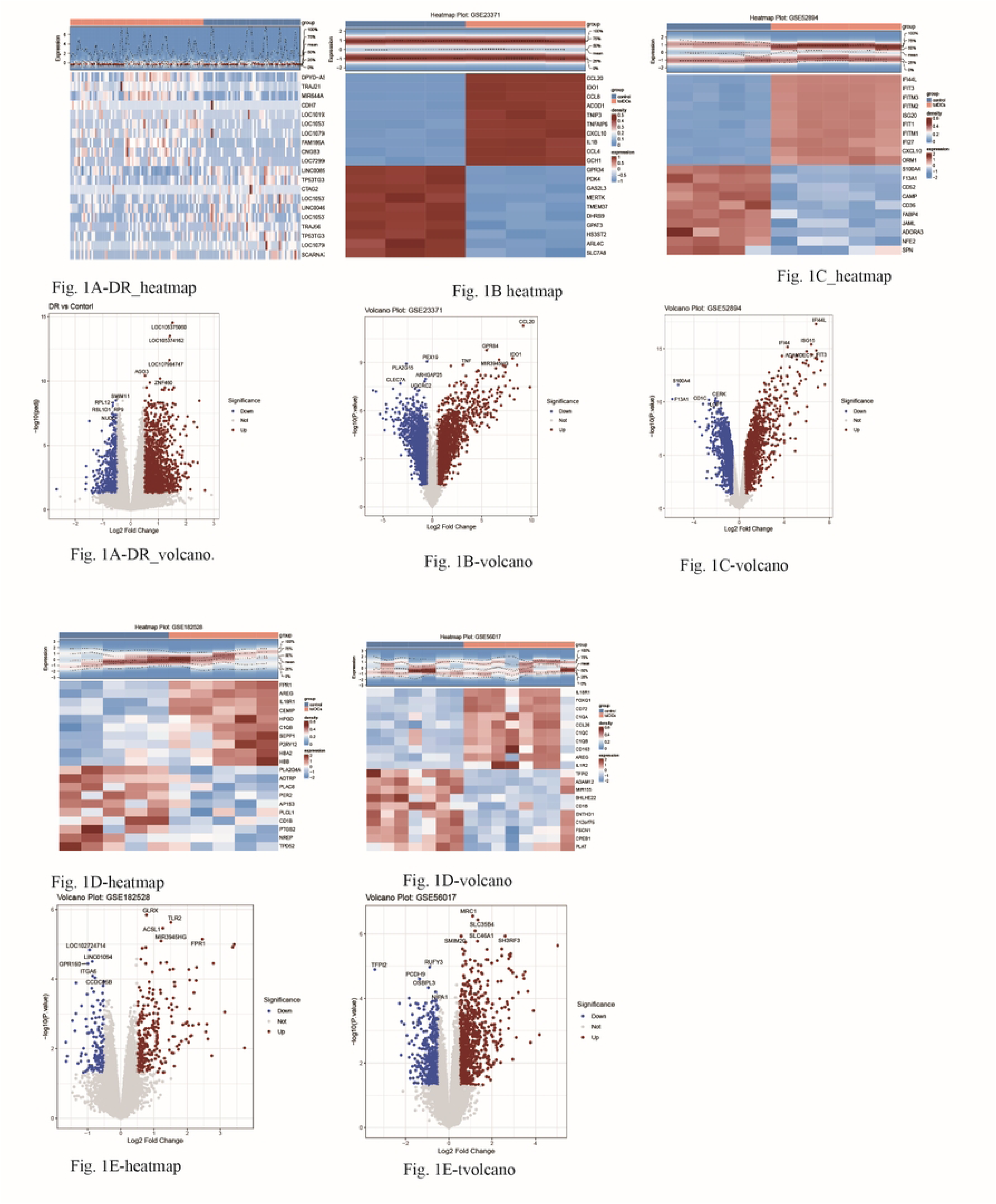
(A) the GSE221521 dataset, (B) the GSE23371 dataset, (C) the GSE52894 dataset, (D) the GSE182528 dataset, (E) the GSE56017 dataset, total 6,267 tolDCs-related genes

### 3.2 Acquisition of 51 key genes

By integrating the tolDCs-related genes with DEGs1, 221 candidate genes were identified (**Fig. 2A**). The identified key genes demonstrated significant enrichment in 386 GO categories and 2 major KEGG signaling pathways. For instance, GO terms included “cytoplasmic translation,” “cytosolic ribosome,” and “cadherin binding” (**Fig. 2B, Supplementary Table 2**). The KEGG pathways primarily included “Coronavirus disease-COVID-19” and “Ribosome” (**Fig. 2C, Supplementary Table 3**). In the PPI network, associations were found between 203 proteins, such as OSA3 and FASN (**Fig. 2D**). A total of 51 key genes were identified by using the 5 algorithms for selection (**Fig. 2E**).

**FIGURE 2.**
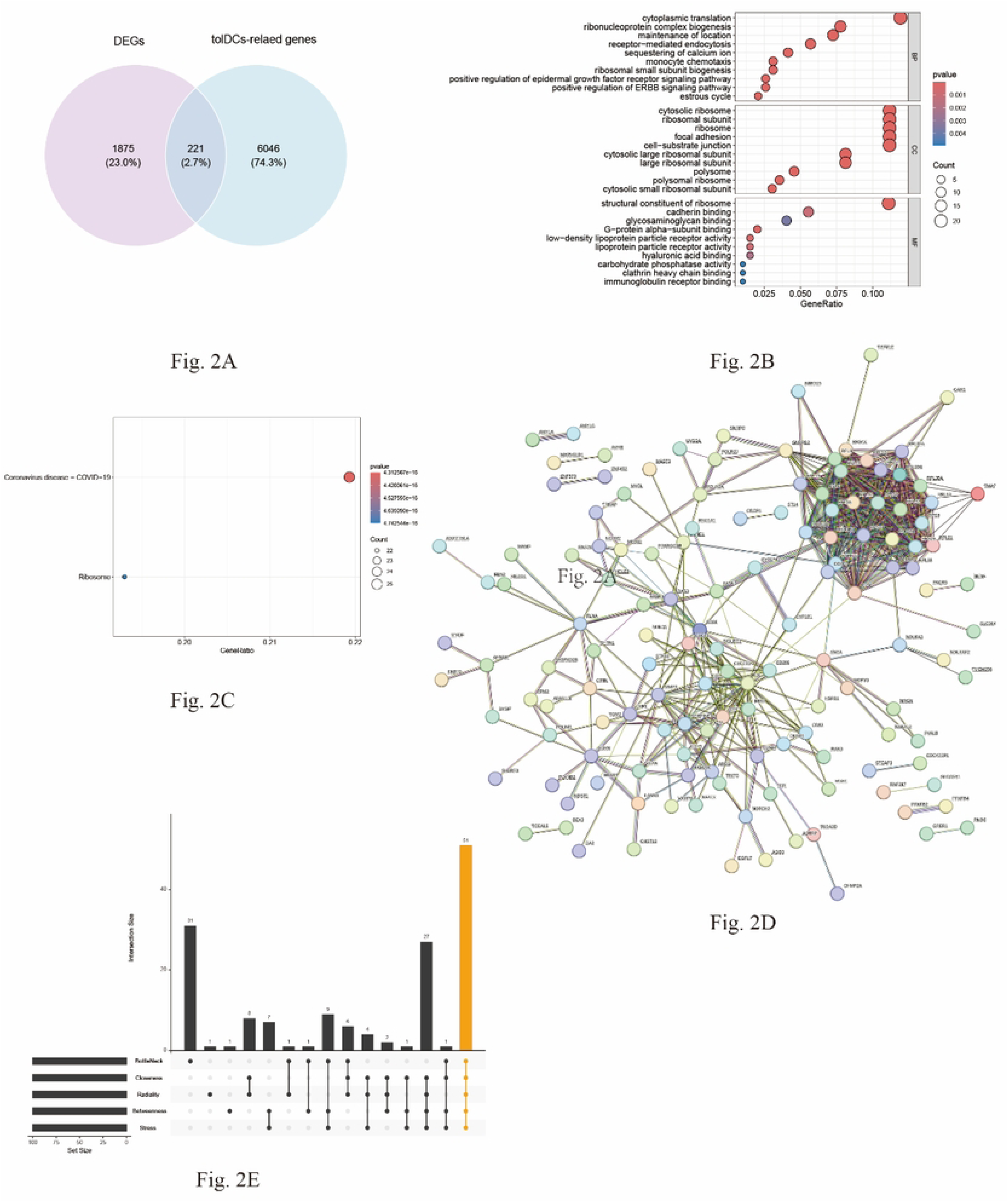
Acquisition of 51 key genes. (A) The Venn diagram shows the candidate genes. (B) The key genes were associated with GO terms. (C) The key genes were associated with 2 KEGG pathways. (D) the PPI network. (E) total of 51 key genes were identified.

### 3.3 Identification of TFEC and CHMP2A as biomarkers

9 feature genes (feature genes 1) were identified using LASSO regression analysis (lambda.min = 0.04) (**Fig. 3A**). Meanwhile, 10 feature genes (feature genes 2) were identified using the Random Forest algorithm (**Fig. 3B**). By integrating feature genes1 and 2, 2 candidate biomarkers were identified: TFEC and CHMP2A (**Fig. 3C**). Cross-dataset validation in GSE221521 and GSE185011 confirmed consistent differential expression patterns, with TFEC demonstrating significant upregulation and CHMP2A showing marked downregulation in DR samples compared to controls. Therefore, TFEC and CHMP2A were classified as biomarkers (*p*< 0.05) (**Fig. 3D**). ROC analysis indicated that both TFEC and CHMP2A had good predictive value for DR, with AUC values of 0.770 and 0.739, respectively (**Fig. 3E**).

**FIGURE 3.**
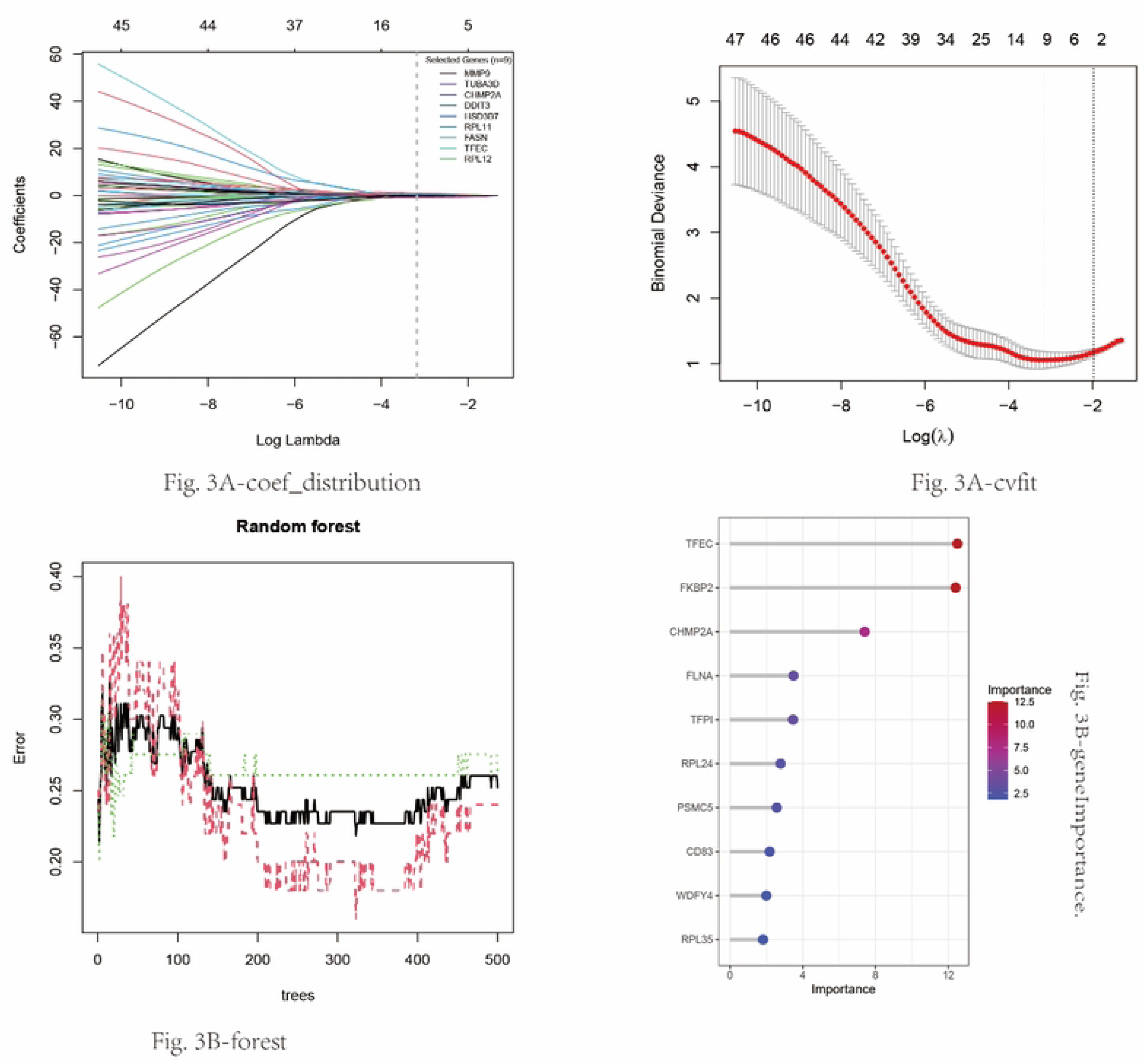
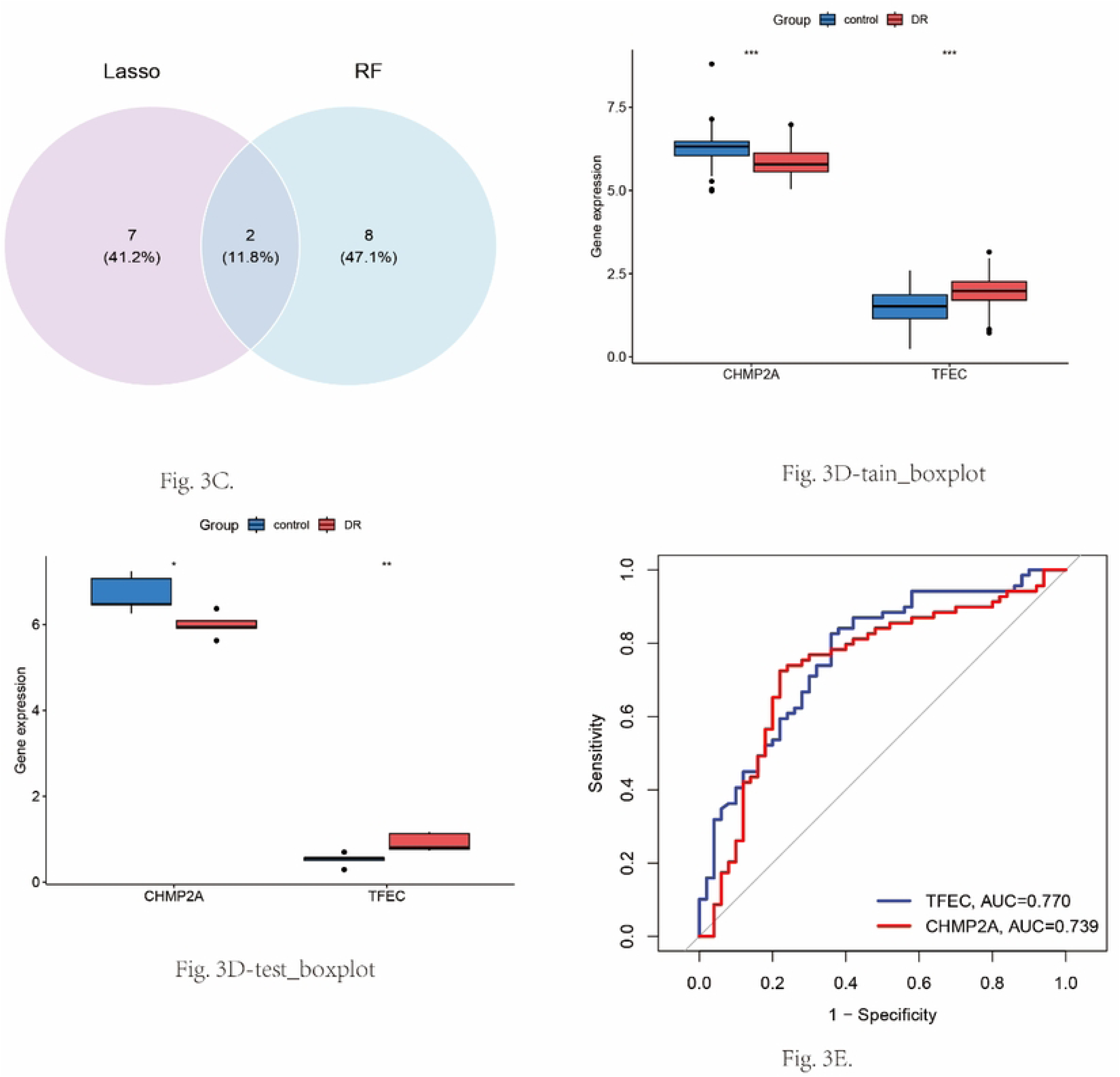
Identification of TFEC and CHMP2A. (A) LASSO regression analysis (lambda min =0.04). (B) RF algorithm. (C) 2 candidate biomarkers-TFEC and CHMP2A. (D) Analysis of candidate biomarkers. (E) ROC analysis.

### 3.4 Multiple signaling pathways in DR

Gene Set Enrichment Analysis (GSEA) identified TFEC as significantly associated with 994 biological pathways (FDR < 0.25), with prominent enrichment in the ’Cytoplasmic Ribosomal Proteins’ pathway. CHMP2A was enriched in 748 pathways, including “tRNA Processing.” The pathways enriched by both TFEC and CHMP2A included “apoptosis,” “VEGFA-VEGFR2 signaling,” and “signaling by VEGF,” among others (**Fig. 4A-B, Supplementary Tables 4-5**). These findings strongly implicate the VEGF-mediated signaling pathway as a critical driver of diabetic retinopathy pathogenesis.

**FIGURE 4.**
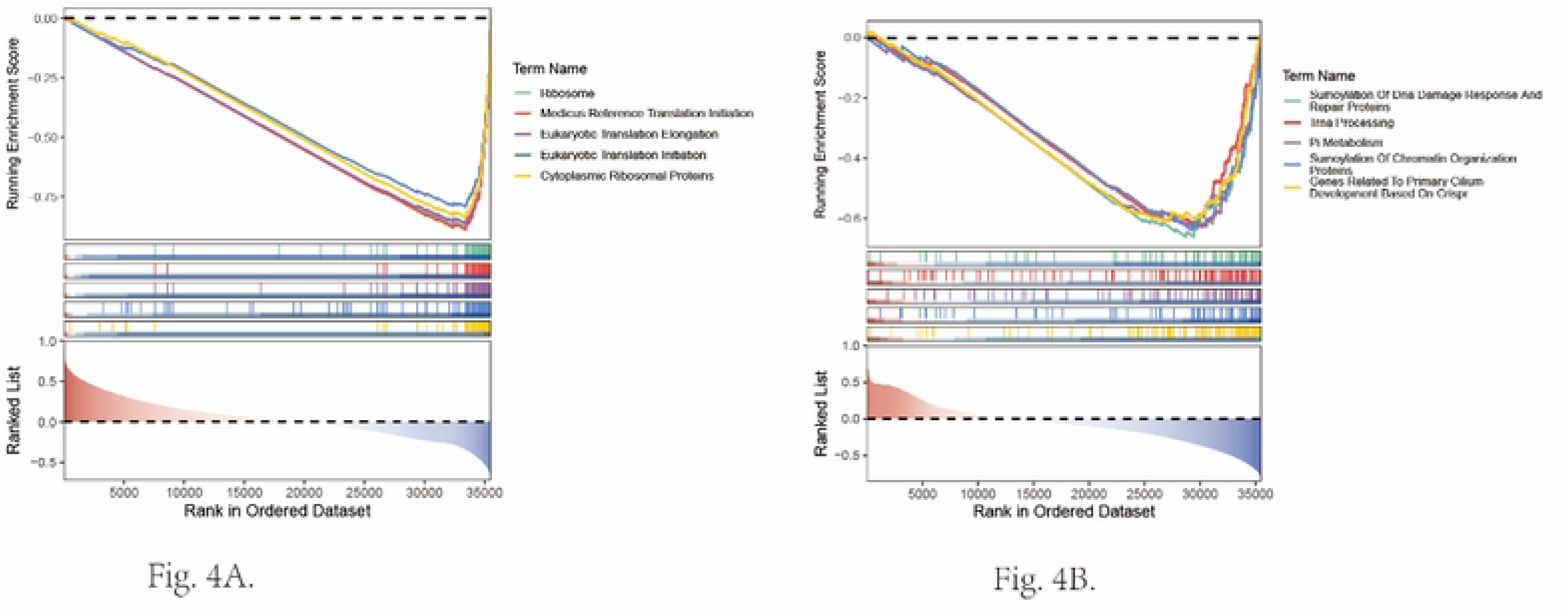

### 3.5 Association of biomarkers with the immune microenvironment

The stacked bar plot demonstrated the percentages of 22 immune cell types in DR and control groups (**Fig. 5A**). The scores of 8 immune cell types showed significant differences, including M0 macrophages (*p* < 0.05) (**Fig. 5B**). Spearman’s correlation analysis demonstrated a significant positive association between activated NK cells and resting mast cells (ρ = 0.46, *p* < 0.001) (**Fig. 5C, Supplementary Table 6**). In addition, Spearman correlation analysis revealed that TFEC showed the strongest correlation with activated NK cells (cor = -0.49, *p* < 0.001), while CHMP2A correlated most strongly with resting mast cells (cor = 0.49, *p* < 0.001) (**Fig. 5D, Supplementary Table 7**). These results indicated that there were interactions between immune cell types and that biomarkers were involved in the progression of DR.

**FIGURE 5.**
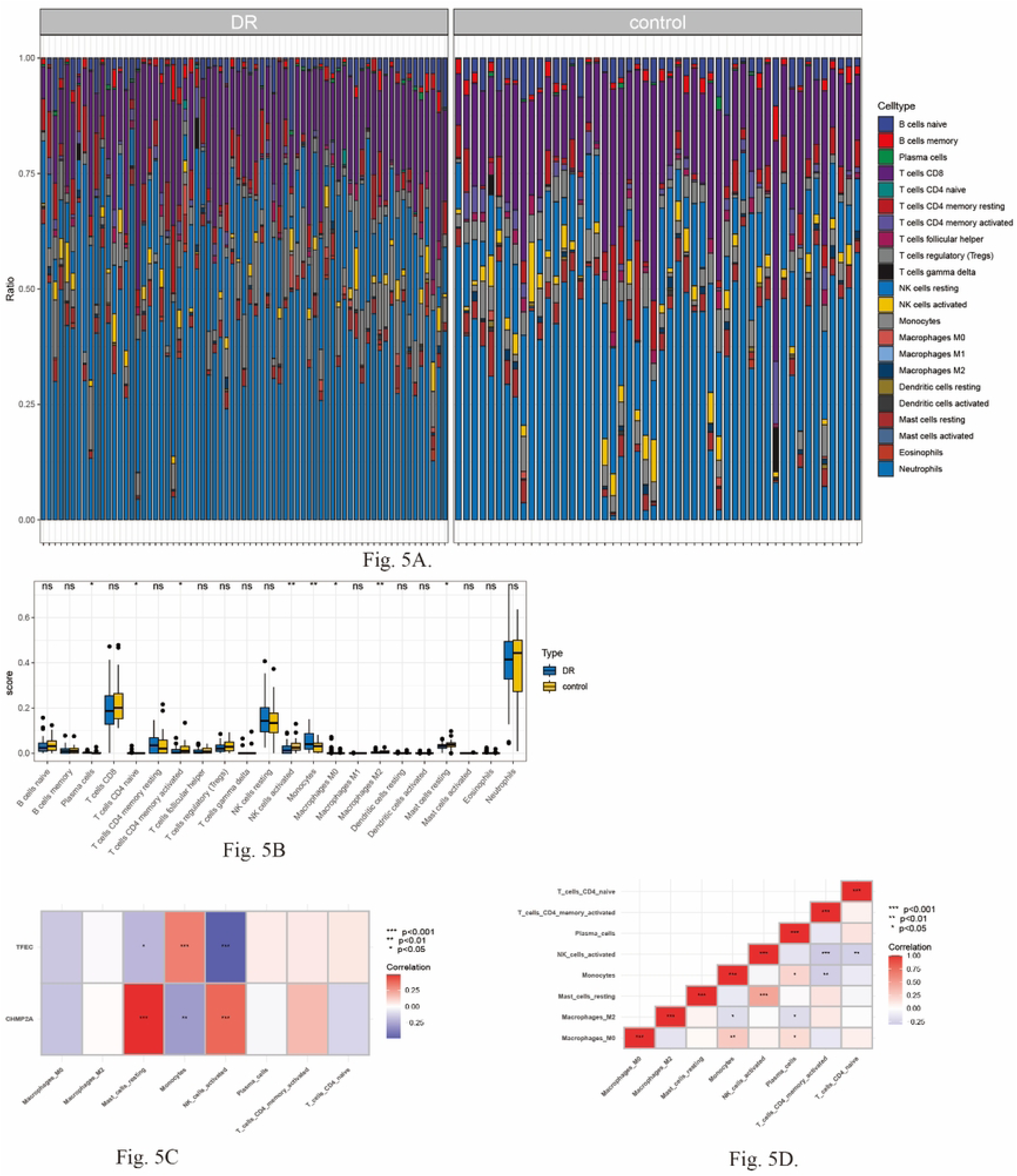
Association of biomarkers with the immune microenvironment. (A) The stacked plot of 22 immune cell types in DR and control groups. (B) scores of 8 immune cell types, MO macrophages (P< 0.05). (C) Spearman correlation analysis activated NK cells and resting mast cells. (D) Spearman correlation analysis TFEC and NK cells.

### 3.6 Clinical experimental results

The cohort consisted of 5 patients diagnosed with DR as the experimental group (3 females and 2 males; mean age, 65.8 ± 8.7 years) and 5 matched subjects as the control group (1 female and 4 males; mean age, 64.8 ± 11.2 years). Statistical analysis showed no significant differences in gender (*p* = 0.524) or age (*p* = 0.880) between the two groups (all *p* > 0.05), indicating comparable baseline characteristics.

RT-qPCR validation confirmed significant upregulation of TFEC (*p* < 0.01) and downregulation of CHMP2A (*p* < 0.05) in DR samples compared to controls, fully concordant with bioinformatics predictions (Fig. 6A-B). This further confirmed the reliability of the bioinformatics analysis.

**FIGURE 6.**
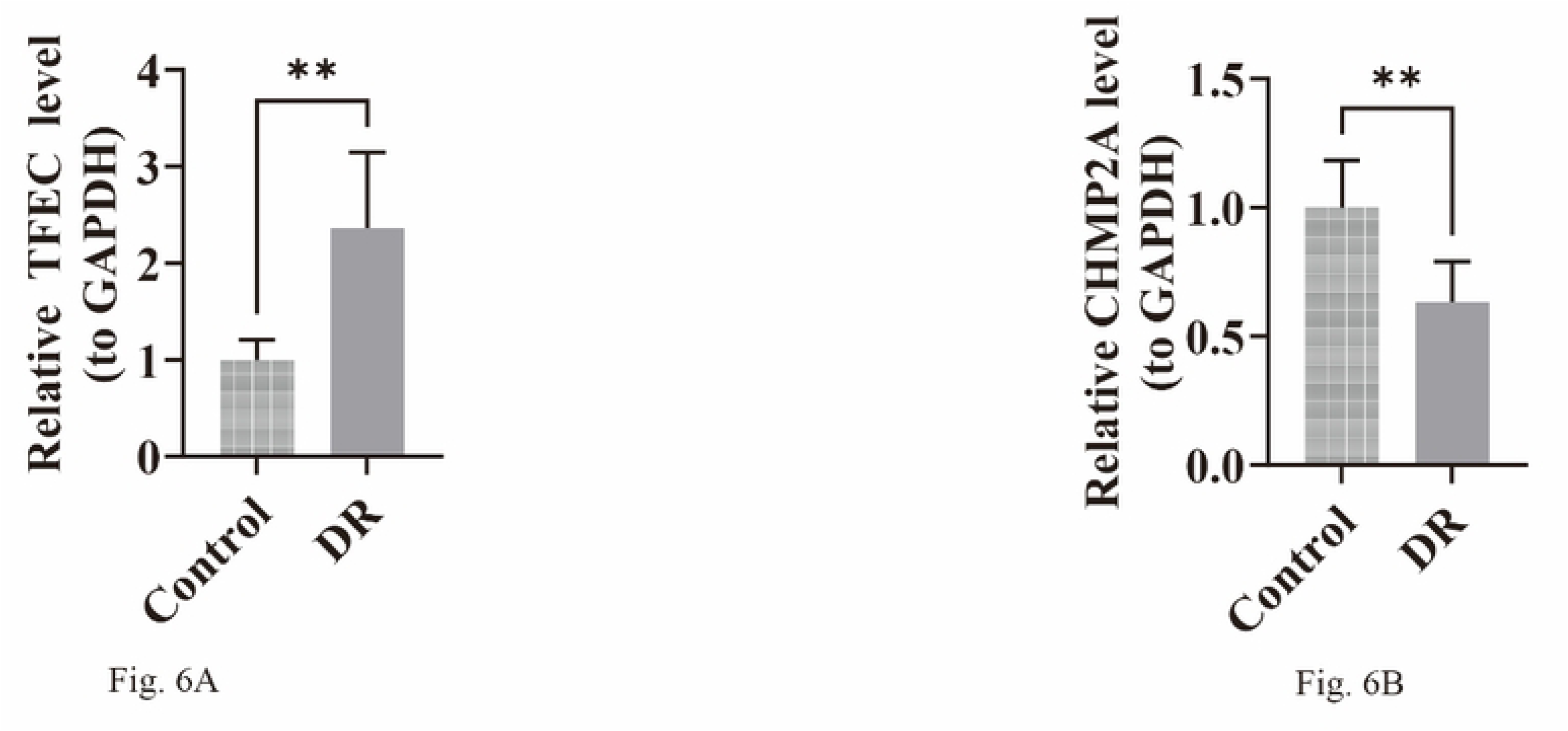
Clinical experimental results. (A-B) RT-qPCR verify TFEC and CHMP2A, P <0.05 indicates statistical significance

## 4. Discussion

DR is a complex clinical disease with multifactorial etiology, and its pathogenesis involves various pathological processes, such as oxidative stress, inflammation and immune dysregulation. Tolerogenic Dendritic Cells (TolDCs) have unique immune regulatory capabilities and play a key role in disease progression [24–26]. This study identified two key genes, TFEC and CHMP2A, using bioinformatics analysis methods based on the GEO database. Further analyses, including GSEA and immune infiltration, established TFEC and CHMP2A as critical regulators in DR pathogenesis, with TFEC promoting and CHMP2A suppressing disease progression, revealing novel therapeutic targets for intervention.

TFEC is a member of the microphthalmia-associated transcription factor (MiT) family, which includes key regulators of melanocyte development and ocular pigmentation. Members of this family act as transcription factors that can regulate the expression of target genes by binding to E-box recognition sequences in the form of homodimers or heterodimers, playing roles in various cellular processes such as cell survival, growth, and differentiation [27]. Experimental data demonstrate significant upregulation of TFEC in cardiac tissue of obese murine models subjected to combined hemodynamic stress and angiotensin II stimulation. Relative to rAd-NC controls, rAd-shTFEC administration markedly decreased TFEC expression in myocardial tissue. Additionally, AngII treatment elevated the expression levels of atrial natriuretic peptide (ANP), brain natriuretic peptide (BNP), and β-myosin heavy chain (β-MHC) in mouse hearts; in contrast to rAd-NC transfection, rAd-sh-TFEC significantly downregulated ANP, BNP, and β-MHC expression. Echocardiography revealed that rAd-sh-TFEC enhanced cardiac function in AngII-challenged mice relative to rAd-NC controls.

TFEC knockout in primary cardiomyocytes reversed AngII-induced cellular hypertrophy. TFEC silencing partially reversed AngII-induced upregulation of ANP, BNP, and β-MHC expression [28]. Existing research suggests TFEC may contribute to cancer progression [29]. However, the function of TFEC in DR was discovered in this study, with GSEA analysis showing that TFEC is enriched in 994 pathways, including “cytoplasmic ribosomal proteins.” The pathways co-enriched with TFEC and CHMP2A include “apoptosis,” “VEGFA-VEGFR2 signaling,” and “VEGF signaling,” among others. These results suggest that VEGF-mediated signaling pathways play a critical role in DR.

CHMP2A (Chromatin modifying protein 2A) is located on human chromosome 19q13.43. As a member of the CHMP family, it forms part of the ESCRT-III complex involved in endosomal sorting. It consists of 222 amino acids and has a molecular weight of approximately 25.1 kDa [30]. Studies have shown that CHMP2A is a gene that can enhance the drug resistance of glioblastoma stem cells (GSCs). CHMP2A induces the death of NK cells by promoting the secretion of extracellular vesicles (EVs) from tumor cells, thereby helping tumor cells evade NK cell-mediated killing. Experimental studies in HNSCC mouse models have demonstrated CHMP2A’s involvement in enhancing tumor cell evasion of NK cell cytotoxicity [31]. Non-healing wounds or diabetic ulcers are serious complications of diabetes, and studies reveal altered CHMP2A protein levels in wound sites of diabetic obese individuals as early as day 1 post-wounding, which is related to the inability to enter the healing proliferative phase [32].

Studies in diabetes have demonstrated that endoplasmic reticulum (ER) stress significantly contributes to the pathogenesis and progression of diabetes and its complications. CHMP2A is involved in the regulation of the endoplasmic reticulum stress response. When endoplasmic reticulum stress occurs, the expression level of CHMP2A changes, and it may regulate the cell’s adaptation and response to endoplasmic reticulum stress by affecting processes such as the sorting, transport of intracellular proteins, and the formation of membrane vesicles. If the function of CHMP2A is abnormal, it may lead to a decreased ability of cells to handle endoplasmic reticulum stress, thereby affecting normal cellular functions and participating in the pathophysiological processes of diabetes and its complications [33]. However, the function of CHMP2A in DR was discovered in this study, in which CHMP2A was enriched in 748 pathways, including “tRNA processing.” The pathways co-enriched with CHMP2A and TFEC include “apoptosis,” “VEGFA-VEGFR2 signaling,” and “VEGF signaling,” among others (Figure 4A-B, Supplementary Tables 4-5). These results indicate that VEGF-mediated signaling pathways play a key role in DR.

Studies demonstrate a significant association between increased circulating VEGF concentrations and development of retinopathy in individuals with type 2 diabetes. The study involved over 800 patients with type 2 diabetes, dividing them into three groups: those without retinopathy, those with early retinopathy, and those with late retinopathy. Comparative analyses were conducted through regular monitoring of serum VEGF levels and fundus examinations. The results indicated that the average VEGF level in patients without retinopathy was 150 pg/ml, while in patients with early retinopathy, the VEGF level increased to approximately 300 pg/ml, and in those with late retinopathy, it further rose to over 500 pg/ml. The VEGF levels were positively correlated with retinopathy severity. When serum VEGF levels exceeded 250 pg/ml, the risk of patients developing retinopathy increased by nearly 70% [34]. The main pathological features of DR include microvascular cell dysfunction, apoptosis, and an imbalance in protein secretion in the extracellular matrix. In high glucose and high oxygen environments, oxidative stress produced by mitochondria can damage retinal neurons and vascular endothelial cells, particularly retinal ganglion cells (RGCs). Apoptosis is closely related to the development of DR [35]. Additionally, IL-1β induces pericyte apoptosis under high glucose conditions through NF-κB activation, thereby increasing endothelial permeability in DR [36].

The pathways co-enriched with TFEC and CHMP2A include “apoptosis,” “VEGFA-VEGFR2 signaling,” and “VEGF signaling,” among others (Figure 4A-B, Supplementary Tables 4-5). These results demonstrate the crucial involvement of VEGF-dependent signaling mechanisms in diabetic retinopathy pathogenesis.

Of the eight immune cell subtypes exhibiting significant variation between DR patients and controls, using M1 macrophages as an example, the local microenvironment in diabetic retinopathy (high blood sugar, hypoxia, oxidative stress, and inflammatory factors) promotes the polarization of macrophages towards M1 macrophages [37], triggering intracellular signaling cascades (including NF-κB activation) [38], increasing M1 phenotype indicators while stimulating pro-inflammatory cytokine release, and worsening retinal inflammation and pathological damage [39]. Detection of specific markers of M1 macrophages in peripheral blood could help with early diagnosis and treatment planning for diabetic retinopathy. Meanwhile, during diabetic retinopathy, high blood sugar and alterations in the local retinal microenvironment can affect the number of regulatory T cells (Tregs) [40, 41]. At first, their numbers go up to help fight inflammation [42], but as the disease progresses to the proliferative stage, their numbers decrease because of ongoing pathological stimulation and cell death, which leads to a drop in the ability to regulate immune homeostasis, making things worse, like inflammation and abnormal blood vessel growth [43],

In clinical samples, Relative to controls, DR samples exhibited significantly elevated TFEC expression and reduced CHMP2A levels. These findings align with the bioinformatics predictions, validating the computational analysis. The concordance between these results helps to link molecular level changes with clinical phenomena, revealing the biological mechanisms of disease occurrence and development, as well as the targets and pathways of drug action.

This study identified two biomarkers related to tolDCs through bioinformatics methods: TFEC and CHMP2A. The biological functions of the biomarkers and their association with the immune microenvironment were analyzed, clinical specimens were analyzed via RT-qPCR to quantify biomarker expression, with results validating the bioinformatics predictions. Nevertheless, this study has limitations, such as a limited sample size and an incomplete elucidation of the biomarkers’ mechanistic roles in DR pathogenesis. Future research should aim to increase the sample size and employ in vitro cell experiments and in vivo animal models to elucidate the molecular mechanisms, to developing more precise and effective treatment strategies. This study gives new ideas for developing treatment strategies for diabetic retinopathy, and future research will keep digging into the causes and treatment targets.

## Data availability statement

The datasets supporting this study are available in public repositories, with repository names and accession codes provided in the manuscript and supplementary files.

## Ethics statement

This research involving human subjects received ethical approval from the Institutional Review Board of Ruikang Hospital Affiliated to Guangxi University of Chinese Medicine (Ethical review approval number: KY2024-213), with all participants providing written informed consent.

## Author contributions

Wenwang Liang, Songman Li, and Chunyi Wei jointly conceived and designed the research protocol. Wenwang Liang and Songman Li are co-first authors of this article, while Chunyi Wei, as the corresponding author, supervised the entire experimental process. During the research, Wenwang Liang, Songman Li, Lingjuan Liu, Junpeng Huang, and Xiaohui Lai performed the experiments; Lijuan Yang collected the clinical diabetic retinopathy samples; and Wenwang Liang and Songman Li conducted the statistical analysis and drafted the manuscript. All authors made substantial contributions and approved the final manuscript.

## Acknowledgments

The authors gratefully acknowledge TCGA for providing open access to their data resources.

